# Metabolic characterization of plasma and cyst fluid from cystic precursors to pancreatic cancer patients reveal metabolic signatures of bacterial infection

**DOI:** 10.1101/2020.11.03.20225524

**Authors:** Ann Morgell, Julie A. Reisz, Zeeshan Ateeb, Haleh Davanian, Susanne E. Reinsbach, Asif Halimi, Rogier Gaiser, Roberto Valente, Urban Arnelo, Marco Del Chiaro, Margaret Sällberg Chen, Angelo D’Alessandro

## Abstract

Pancreatic cancer is the seventh leading cause of cancer-related death worldwide, with a 5-year survival rate as low as 9%. One factor complicating the management of pancreatic cancer is the lack of reliable tools for early diagnosis. While up to 50% of the adult population has been shown to develop precancerous pancreatic cysts, limited and insufficient approaches are currently available to determine whether a cyst is going to progress into pancreatic cancer. Recently, we used metabolomics approaches to identify candidate markers of disease progression in patients diagnosed with intraductal papillary mucinous neoplasms (IPMNs) undergoing pancreatic resection. Here we enrolled an independent cohort to verify the candidate markers from our previous study with orthogonal quantitative methods in plasma and cyst fluid from serous cystic neoplasm and IPMN (either low- or high-grade dysplasia or pancreatic ductal adenocarcinoma). We thus validated these markers with absolute quantitative methods through the auxilium of stable isotope-labelled internal standards in a new independent cohort. Finally, we identified novel markers of IPMN status and disease progression – including amino acids, carboxylic acids, conjugated bile acids, free and carnitine-conjugated fatty acids, purine oxidation products and TMAO. We show that the levels of these metabolites of potential bacterial origin correlated with the degree of bacterial enrichment in the cyst, as determined by 16S RNA. Overall, our findings are interesting per se, owing to the validation of previous markers and identification of novel small molecule signatures of IPMN and disease progression. In addition, our findings further fuel the provoking debate as to whether bacterial infections may represent an etiological contributor to the development and severity of the disease in pancreatic cancer, in like fashion to other cancers (e.g., *Helicobacter pylori* and gastric cancer).

**Key points:**

- We identified and quantified novel markers of IPMN cyst status and pancreatic cancer disease progression – including amino acids, carboxylic acids, conjugated bile acids, free and carnitine-conjugated fatty acids, purine oxidation products and TMAO.
- We show that the levels of these metabolites of potential bacterial origin correlated with the degree of bacterial enrichment in the cyst, as determined by 16S RNA.

## Introduction

Pancreatic cancer (PC) is the seventh leading cause for cancer related death worldwide (1). Incidence and mortality rates are almost identical (458,918 new cases and 432,242 deaths in 2018) and although improvements with adjuvant chemotherapy have been made, 5-year survival rates remain as low as 9% (1, 2). Incidence has shown an increasing tendency, and PC is estimated to become the second leading cause of cancer-related death by 2030 (3).

There are to this day no effective screening programs where pre-cancerous lesions can be detected with satisfactory accuracy and treated accordingly. Much research today therefore aims to improve diagnostic measures of pre-cancerous cystic lesions of the pancreas. The main proportion of these pancreatic cystic neoplasms (PCN) consists of intraductal papillary mucinous neoplasms (IPMN), which have a potential to transform from low grade dysplasia (LGD) through high-grade dysplasia (HGD) to cancer. Discriminating these potentially malignant cysts from essentially benign cysts of the pancreas, such as serous cystic neoplasms (SCN) is a delicate matter. A few years ago, pre-operative diagnostic accuracy was only around 60-70% even in expert centres (4). Adding endoscopic ultrasound and fine needle aspiration (EUS-FNA) adds some diagnostic information, as does blood levels of carcinoembryonic antigen (CEA), but they do not provide satisfactory information on the grade of dysplasia in these lesions, and cytology is not sufficiently specific to distinguish a mucinous lesion with LGD from one that has already transformed into cancer (5–7). Though diagnostic accuracy is continuously improved, including early recognition of main duct dilation (8), discriminating and grading these potentially pre-malignant cysts is still challenging.

The prevalence of cystic lesions in the pancreas is very high in the general population, up to 49.1% as some studies indicate (9) – with an average number of ∼4 cysts per subject (9). The incidence of pancreatic cancer, however is estimated to be 5.5 and 4.0 per 100,000 in men and women, respectively, worldwide (10) and, indeed, a 5 year follow-up of 1077 subjects with ∼50% pancreatic cyst incidence observed no pancreatic cancer (9). This means that a small minority of these cysts will eventually evolve to cancer, adding further to the delicate dilemma of how to handle these cysts.

Since pancreas resection involves high risk surgery, with significant peri-operative morbidity and mortality, optimising pre-operative diagnostics is of outmost importance in order to select the patients that truly will benefit from surgery from those that will not. Recent studies from our group indicates that by integrating metabolomics and lipidomics approach it is possible to discriminate IPMN from SCN, and the grade of dysplasia associated with IPMN, with high sensitivity and specificity (11). In spite of a limited sample size and a workflow optimized for lipidomics characterization, we found that major key discriminating compounds were sugars, nucleotides or amino acids. While promising, our early study did not manage to discriminate IPMNs with different grade of dysplasia from pancreatic cancer. The lack of discriminating power of the data generated in the previous study could be potentially overcome by leveraging untargeted metabolomics approaches and the comprehensive output they generate, as is in part the focus of this manuscript. In addition, candidate markers of IPMN status progression that were identified in our previous study had not been quantified beyond the determination of relative measurements (peak areas in arbitrary units). In the present study, we aim to validate those results using an independent metabolomic platform on a new set of cyst fluid and plasma samples collected from a Swedish cohort undergoing pancreas resection due to suspicion of IPMN.

Moreover, since publication of our first metabolomics report, we have reported the detection of an intracystic pancreatic microbiome, as gleaned by 16S RNA gene sequencing analysis (12). Here we consolidate those findings and report a significant correlation between IPMN dysplasia or cancerous state and plasma/cyst fluid levels of metabolites of potential bacterial origin such as trimethylamine-oxide (TMAO), polyamines, carboxylic acids and several conjugated bile acids.

## Results

### Metabolomics analyses of plasma and cyst fluid from SCN, IPMN and PDAC patients

The patients’ population is extensively described in **Table 1**. All patients diagnosed with SCN were women, which is expected as SCN is present in women to a much larger extent than in men (13). As expected, the SCN groups were also younger than the other groups. As for LGD, HGD and PDAC, they were rather evenly distributed between the sexes. Commonly used blood parameters (HbA1c, S-Ca 19-9, S-amylase, albumin, bilirubin, and white blood cell count) didn’t vary across the groups, except for S-Ca 19-9 and bilirubin levels, which were both significantly higher in the PDAC group where plasma samples were retrieved. To expand on previous targeted metabolomics analyses on plasma and cyst fluid of patients presenting with IPMN or PDAC (11), untargeted metabolomics analyses were performed on plasma (**Figure 1**) and cyst fluid (**Figure 2**) from an independent validation cohort, enrolled at the Karolinska Institute in Stockholm. Plasma analyses (**Figure 1.A**) identified 38 unique compounds/chemical formulae increasing in plasma from SCN patients compared to PDAC patients and 70 significantly following an opposite trend (**Figure 1B**). A series of representative volcano plot report similar analyses comparing HGD to PDAC, HGD to LGD and LGD to PDAC, the latter showing the largest differences (1433 unique chemicals increasing in LGD and 782 decreasing compared to PDAC – **Figure 1.B**). Pathway analyses of these chemicals highlighted a significant impact of disease progression on amino acid metabolism (**Figure 1.C**), carboxylic acids (**Figure 1.D**) and glycolysis-related metabolites (glucose and lactate, substrate and product of the pathway, respectively - **Figure 1.E**), confirming and expanding upon previous reports (11).

**Table 1.** Patient group characteristics.

|  | Cyst fluid (n = 57) |  |  |  | Plasma (n = 45) |  |  |  |
| --- | --- | --- | --- | --- | --- | --- | --- | --- |
|  | SCN | IPMN |  |  | SCN | IPMN |  |  |
|  |  | LGD | HGD | Cancer |  | LGD | HGD | Cancer |
| Patients, % (n) | 8.8 (5) | 50.9 (29) | 14.0 (8) | 26.3 (15) | 11.1 (5) | 44.4 (20) | 22.2 (10) | 22.2 (10) |
| Female, % | 100 | 59 | 62 | 53 | 100 | 50 | 70 | 40 |
| Alcohol use, % | - | - | - | - | - | - | - | - |
| Smokers, % | 0 | 19 | 13 | 29 | 0 | 15 | 0 | 20 |
| CVD, % | 40 | 62 | 75 | 53 | 40 | 60 | 70 | 60 |
| Statins use, % | 0 | 21 | 25 | 13 | 20 | 21 | 40 | 20 |
| Diabetes, % | 0 | 21 | 50 | 27 | 0 | 20 | 60 | 20 |
| Age, years | 47 | 70*** | 76*** | 71*** | 47 | 68*** | 74*** | 76*** |
| median (range) | (30-68) | (47-82) | (67-84) | (55-82) | (30-68) | (55-79) | (66-84) | (65-83) |
| BMI, kg/m<sup>2</sup> | 33.69 | 26.39** | 28.56 | 24.21*** | 28.01 | 26.87 | 28.04 | 24.08 |
| median (range) | (21.6-37.1) | (20.8-40.4) | (23.5-32.0) | (18.8-30.2) | (21.6-37.1) | (21.6-36.7) | (23.4-30.5) | (18.8-29.5) |
| HbA1c, mmol/mol | 40 | 40 | 52 | 41 | 41 | 41 | 42 | 40 |
| median (range) | (33-49) | (32-58) | (28-68) | (15.1-60) | (36-49) | (32-62) | (28-68) | (31-67) |
| S-CA 19-9, kE/L | 9.9 | 9.4 | 24 | 39 | 7.9 | 11 | 24 | 255*** |
| median (range) | (<3-34) | (<1-60) | (9-71) | (<1-4840) | (<3-11) | (<1-182) | (<1-189) | (<1-694) |
| Serum amylase, μkat/L | 0.46 | 0.43 | 0.53 | 0.6 | 0.46 | 0.44 | 0.27 | 0.6 |
| median (range) | (0.3-0.75) | (<0.13-1.45) | (<0.13-4.18) | (<0.13-11.6) | (0.19-0.75) | (<0.13-1.45) | (<0.13-4.18) | (<0.13-2.07) |
| Albumin, g/L | 37 | 37 | 36 | 34 | 37 | 36 | 36 | 34 |
| median (range) | (33-40) | (31-42) | (16-39) | (22-38) | (33-40) | (31-42) | (16-39) | (22-38) |
| Bilirubin, μmol/L | 5 | 6 | 8 | 8 | 5 | 6.8 | 5 | 19*** |
| median (range) | (5-9) | (<3-34) | (<3-12) | (4-127) | (3-9) | (3-34) | (3-25) | (4-119) |
| WBC, x 10<sup>9</sup>/L | 7.6 | 7.0 | 7.6 | 7.9 | 7.1 | 7.1 | 8.4 | 10.2 |
| median (range) | (4.1-10.9) | (0.3-11.3) | (5.5-9.4) | (4.2-10.8) | (4.1-10.9) | (0.3-11.3) | (6.4-12.9) | (4.2-13.9) |

Statistical comparisons between each group and the control group (SCN) were made using Stata 15 software and quantile regression for all quantitative measures; \* $p \leq 0.05$ , \*\* $p \leq 0.01$ , \*\*\* $p \leq 0.001$ .

**Figure 1.**
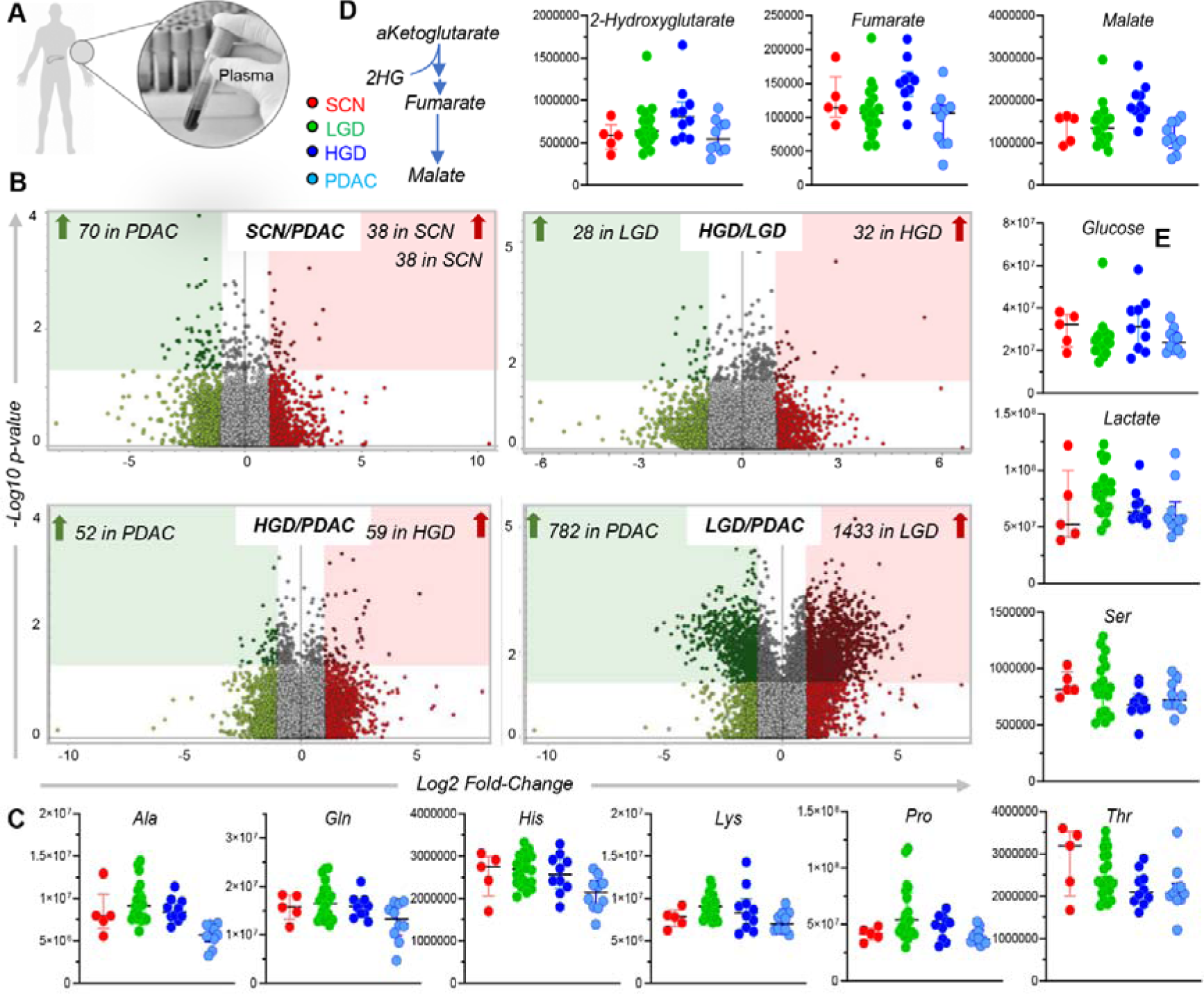
Untargeted metabolomics analyses of plasma. **(A) revealed distinct signatures in IPMN and PDAC patients**, as shown in the volcano plots from the untargeted metabolomics analyses (**B**). Top pathways included amino acid metabolism (**C**), carboxylic acids (**D**) and glycolysis-related metabolites (glucose and lactate, substrate and product of the pathway, respectively - **E**).

**Figure 2.**
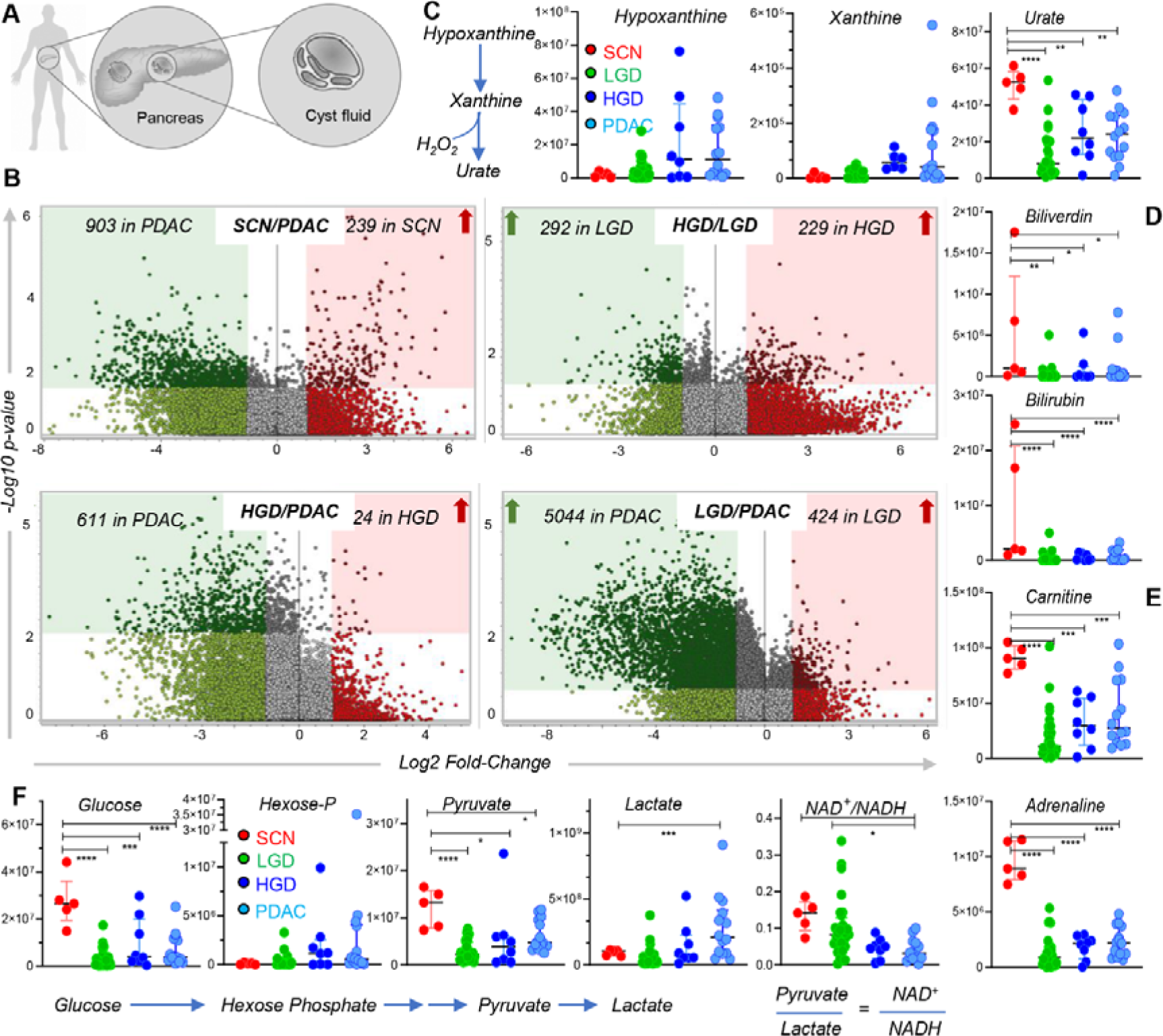
Untargeted metabolomics analyses of cyst fluid. **(A) revealed distinct signatures in IPMN and PDAC patients**, as shown in the volcano plots from the untargeted metabolomics analyses (**B**). Top pathways included purine oxidation (**C**), heme metabolism (**D**) and acyl-carnitines (**E**). glycolytic metabolites (**F**),

Similar untargeted metabolomics analyses were performed on cyst fluid (**Figure 2**). Volcano plots in **Figure 2.B** highlight a significant divergent small molecule composition across the four groups – with phenotypes more markedly distinct across the groups than those observed for plasma. From this analysis, top pathways identified across the four groups included purine oxidation (hypoxanthine, xanthine and urate - **Figure 2.C**), heme metabolism (biliverdin and bilirubin - **Figure 2.D**), acyl-carnitines (*see next paragraphs -* **Figure 2.E**), and glycolytic metabolites (glucose, hexose phosphate isobars, pyruvate and lactate - **Figure 2.F**). To validate untargeted metabolomics analyses, additional semi-targeted metabolomics analyses were performed (untargeted acquisition, post-hoc manual validation of high-resolution accurate intact mass, isotopic patterns, chemical formula composition and retention times against an in-house library of pure chemical standards). A total of 123 metabolites in plasma and 161 in cyst fluid were manually validated through targeted metabolomics analyses – **Supplementary Table 1**). Multivariate analyses of targeted metabolomics data are presented in **Figure 3.B** and **C**, which show the results from partial least square-discriminant analysis (PLS-DA) and hierarchical clustering analysis (top 25 metabolites by ANOVA p-values) in plasma and cyst fluid, respectively. Hierarchical clustering analysis, ANOVA and PLS-DA of semi-targeted data highlighted similar changes to untargeted metabolomics in top metabolic pathways (e.g., carboxylic acids, amino acids, glycolytic metabolites and purine oxidation products changed significantly in plasma – especially in HGD patients – **Supplementary Figure 1**). This analysis was necessary to verify the chemicals identified as significant in the exploratory, untargeted metabolomics analyses. These results were further validated with orthogonal, quantitative methods against stable isotope-labelled internal standards.

**Figure 3.**
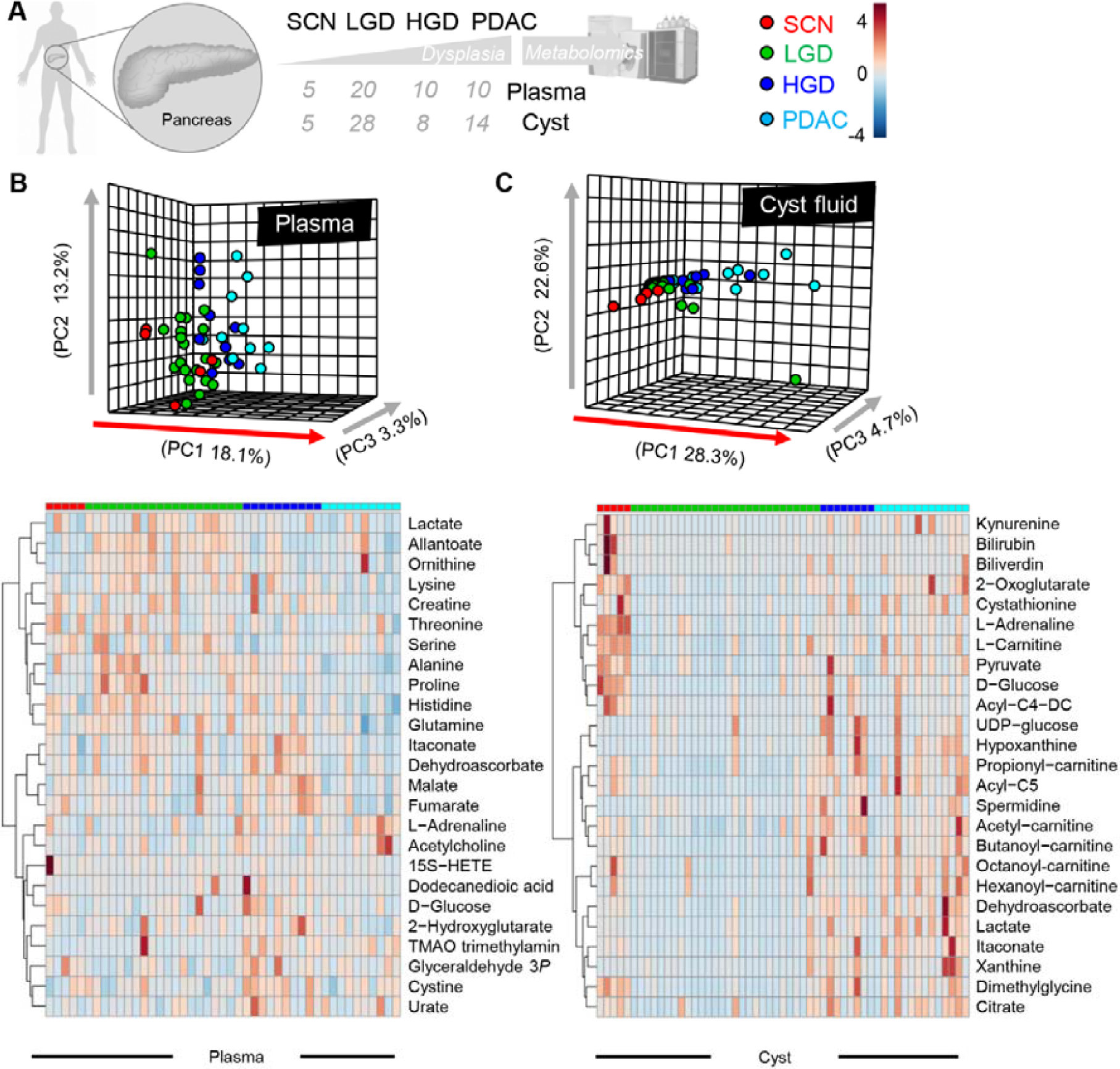
Metabolomics analyses of plasma and cyst fluid from IPMN and PDAC patients. An overview of the experimental design is provided in **A**. in **B** and **C**, results from partial least square-discriminant analysis (PLS-DA) and hierarchical clustering analysis (top 25 metabolites by ANOVA p-values) in plasma and cyst fluid, respectively.

### Biomarker analysis in plasma and cyst fluid based on absolute quantitative measurements of top metabolites from the untargeted and targeted analyses

Absolute quantitation against internal standards was performed for the most significant metabolites from the exploratory metabolomics analyses. Absolute quantitative data were leveraged to determine specificity and sensitivity of top biomarkers of pre-cancerous and cancerous (HGD or PDAC) vs non-cancerous IPMN and serous cystic neoplasm (LGD or SCN) in cyst-fluid and plasma. Results from these analyses are provided as part of **Supplementary Table 1**, including ranked biomarkers as a function of Areas Under the Curve (AUC), p-values, confidence interval and thresholds. Analysis of cyst fluid (**Figure 4.A**) identified amino acids (especially choline – Area Under the Curve, AUC: 0.78 at 7.06 uM), carboxylic acids (succinate, fumarate and malate, all with AUC: >0.75) and several acyl-carnitines (AUC > 0.75 for 9 different carnitines) amongst the top biomarker of cancerous state (**Figure 4.B**). A detail of absolute carnitine concentrations in cyst fluids is shown in **Figure 4.C**, highlighting this class of compounds as the top discriminants between non-cancerous (SCN and LGD) and IPMN cysts showing HGD or PDAC association. Of note, acyl-carnitines ranked amongst the top markers of PDAC when comparing absolute quantitative measurements of cyst fluid from this group to all other groups (SCN, LGD and HGD combined – **Supplementary Figure 2**). On the other hand, biomarker analyses of cyst fluid from SCN vs PDAC patients identified the following top biomarkers: amino acids (glutamine, glutamate, adrenaline, valine, leucine, kynurenine), sugars (glucose, ribose), gamma-glutamyl-cycle metabolites (glutamyl-selenomethionine and aminobutyrate, 5-oxoproline), purine oxidation products (hypoxanthine, allantoate) and carboxylic acids (fumarate - **Supplementary Figure 3**). Carboxylic acids (malate), aromatic amino acids and related metabolites (e.g., tryptophan, indole, dopamine, adrenaline) were identified as the top discriminant between cyst fluid of HGD and PDAC patients (**Supplementary Figure 4**). Similar analyses in plasma (**Figure 4.D**) highlighted the bacterial metabolite trimethylamine-oxide (TMAO) as the top biomarker (AUC: 0.82 at concentration 9.12 uM). Similarly, several conjugated bile acids were identified amongst the top plasma markers of cancerous IPMN, above all taurochenodeoxycholate (AUC: 0.73, 204 nM concentration) (**Figure 4.E**). Indeed, 5 conjugated bile acids ranked amongst the 7 top discriminants between cyst fluid from PDAC patients versus any other cyst (SCN, LGD and HGD – **Supplementary Figure 2**). Notably, dysregulation of bile acid metabolism has been reported as a marker of bacterial metabolism (e.g., in the context of inflammation, obesity, haemorrhagic shock or iron-induced dysbiosis (14–17)), since bile acids can be deconjugated by bacteria.

**Figure 4.**
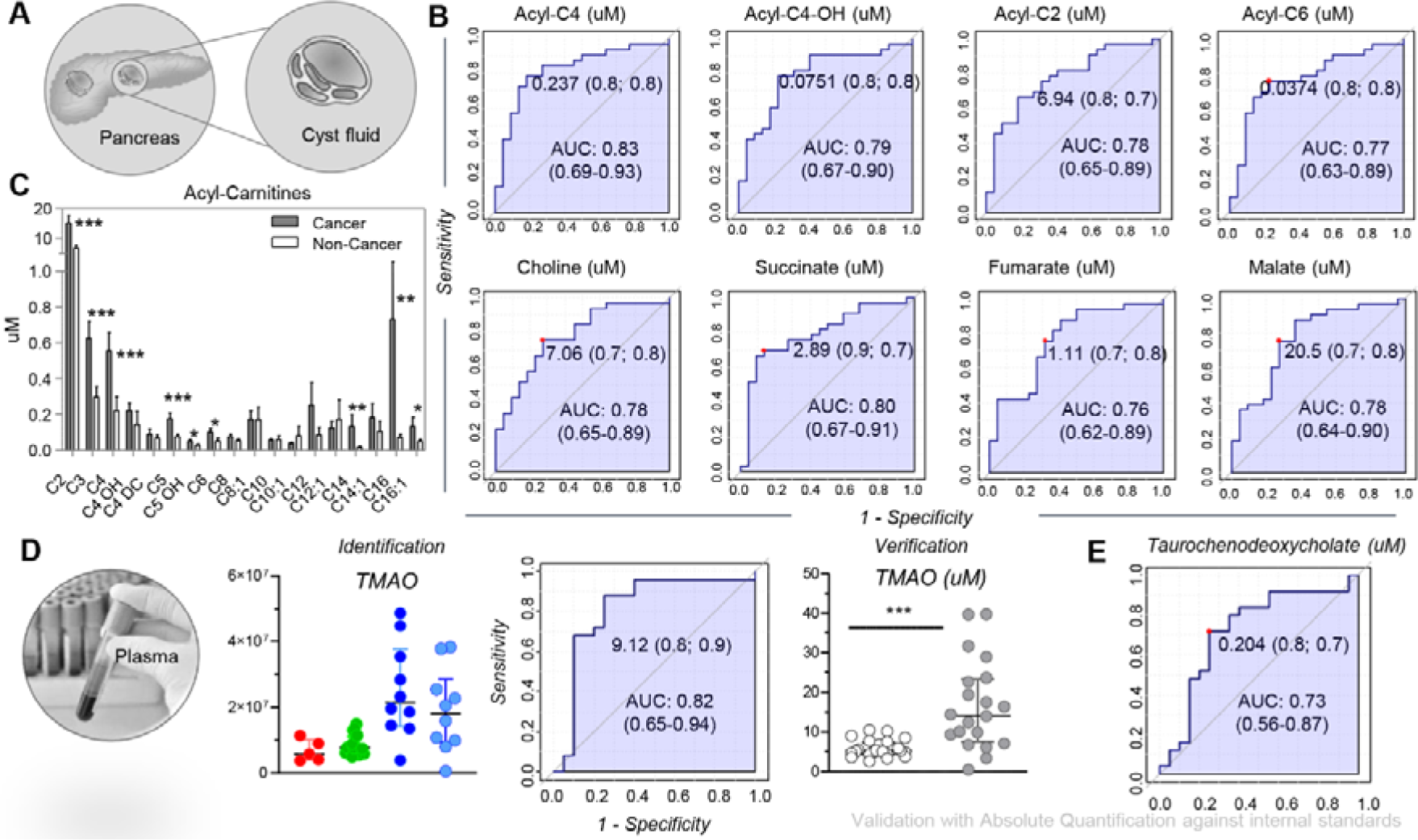
Absolute quantitation of the most significant metabolites from the exploratory metabolomics analyses were leveraged to determine specificity and sensitivity of top biomarkers in cyst-fluid and plasma. Analysis of cyst fluid (**A**) identified amino acids, carboxylic acids and – above all – acyl-carnitines (ROC curves in **B**, bar plots in **C**) as the top discriminants between non-cancerous IPMN (SCN and LGD) and cysts showing HGD or derived from PDAC patients. Similar analyses in plasma (**D**) highlighted the bacterial metabolite trimethylamine-oxide (TMAO) as the top biomarker, an observation validated with orthogonal quantitative methods (uM concentration provide in the ROC curve and dot plot in **D**). Similarly, several conjugated bile acids – that are deconjugated by bacteria in humans – were identified amongst the top plasma markers of cancerous IPMN (**E**).

### 16S rRNA gene analysis of IPMN cyst fluid (A) revealed the presence a substantial bacterial diversity in the cysts from LGD, HGD and PDAC patients

Recently, we used 16S rRNA gene sequencing analysis to provide evidence documenting bacterial involvement in cysts from patients suffering from IPMN (12). Given the metabolic signature potentially associated with microbial metabolism in the cyst fluid and plasma of patients with HGD and PDAC (**Figure 4**), here we correlate 16S gene diversity of cyst fluid from these patients in which the 16S gene was detectable in cysts from LGD and HGD IPMN, as well as PDAC patients (N=17) (**Figure 5.A**). Analysis of 16S gene sequences revealed a wealth of bacterial families in the cysts, with the most common belonging to the phylum class of *Firmicutes, Proteobacteria, Actinobacteria* and *Alphaproteobacteria* (**Figure 5.B**). Specifically, *Methylobacterium, Klebsiella* (both Gram negative *Proteobacteria*) and *Enterococcus* (Gram positive *– Firmicutes*) ranked amongst the top three most abundant genus members in the cysts tested in this study (**Supplementary Figure 5 –** which also included the top 25 most common bacteria by 16S RNA identified here).

**Figure 5.**
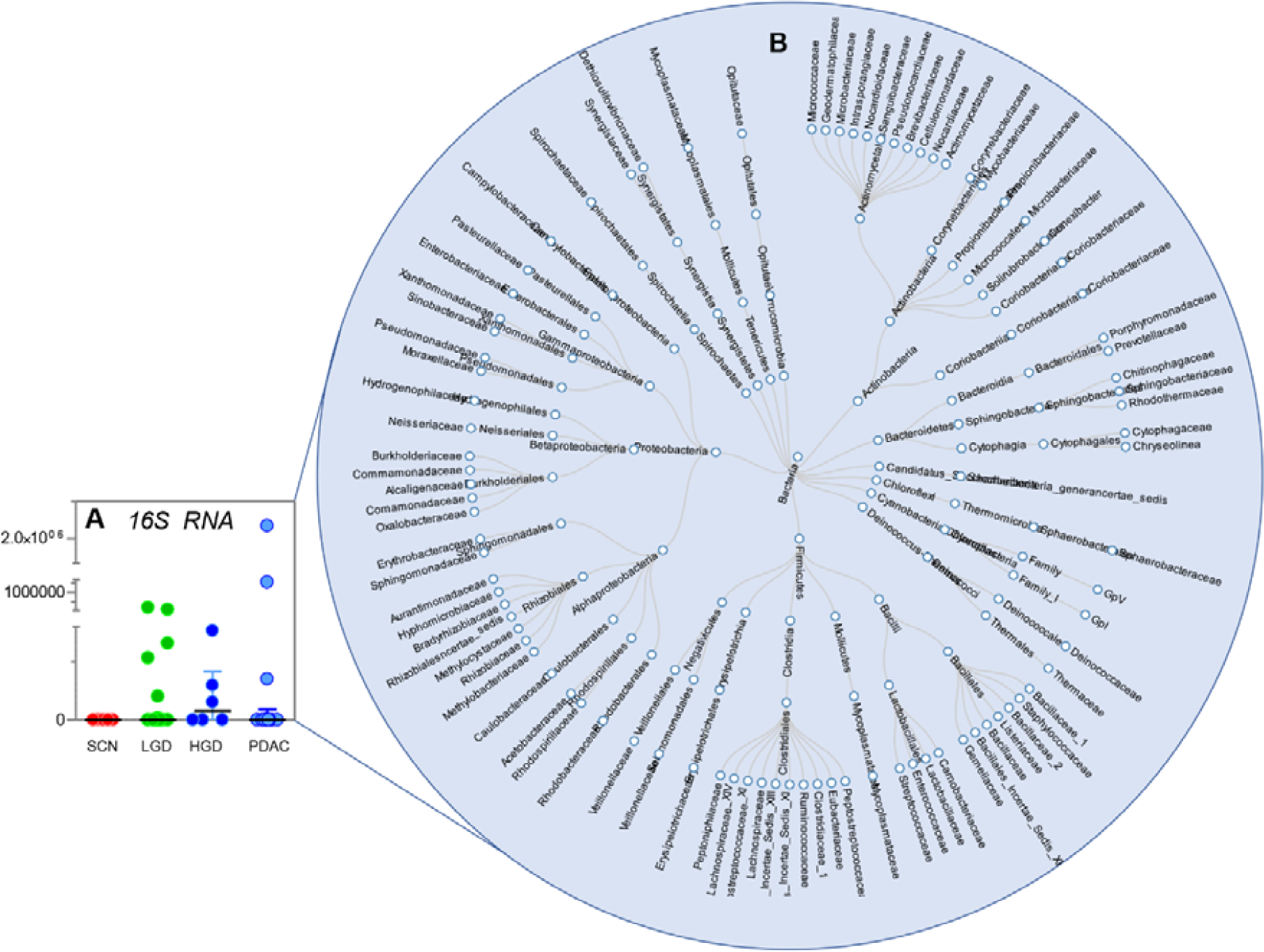
16S RNA gene analysis of IPMN cyst fluid revealed the presence a large bacterial diversity in the cysts from LGD, HGD and PDAC patients (B), with median 16S rRNA gene copy levels indicated from LGD to HGD and PDAC. Each dot represents one patient (A).

Correlation of the cyst 16S gene copy numbers and metabolite levels (**Figure 6.A**) further highlighted a strong correlation (Spearman r > 0.95) for several bile acids (taurolithocholate, deoxycholate, chenodeoxycholate, ursodeoxycholate, cholate, taurodeoxycholate, glycochenodeoxycholate, taurochenodeoxycholate, taurocholate), odd-chain and oxidized fatty acids (C13 and carnitine conjugated-C13, 9(10)diHoME), carboxylic acids (succinate) and arginine/polyamine metabolites (citrulline, cadaverine, putrescine – **Figure 6.B**). Representative correlation curves are shown for several bile acids and succinate, showing that most of these correlations were driven by a subset of PDAC and HGD samples with significantly higher levels of these metabolites and total 16S gene copy number (up to two orders of magnitude for both) compared to the rest of the samples – **Figure 6.C**). Overall, here we identified and quantified metabolites of potential bacterial origin in cyst fluid that was also positive for bacterial infiltration, as determined by 16S gene detection (**Figure 6.D**). Expanding on the intriguing observations that 16S gene copy number in cyst fluid correlated with a subset of metabolites, we correlated metabolite levels to the pancreatic cyst microbiome profiles in the 16S gene sequenced samples. These analyses revealed clusters of metabolites and bacterial phyla to genus level (**Supplementary Table 2**). A hierarchical clustering analysis supported by Spearman correlation coefficient showing the correlations of top significant metabolic and specific bacteria at genus level is provided in **Supplementary Figure 6** (a vectorial version of this heat map is provided in **Supplementary Figure 7**). A hierarchical clustering analysis highlighting the top metabolic correlates to specific bacterial strains is provided as **Figure 7.A** (expanded vectorial version in **Supplementary Figure 8**), while a few selected line plots for top correlates (though no clear trend with respect to the grade of dysplasia was noted) are shown in **Figure 7.B** (extended in **Supplementary Figure 9)**. Overall, these analyses sub-classified two main clusters that involved mainly *Proteobacteri* and *Actinobacteria*. Interestingly, the bile acids and the microbiota-derived metabolites TMAO, cadaverine, putrescine, as well as choline and carnitine which are involved in conversion of TMAO, and the tryptophan pathway derived indole and kynurenine, were mostly found positively correlated with the bacteria in cluster 2. Included this cluster were *Enterobacteriacea, Granulicatella, Klebsiella. Stenotrophomonas, Streptococcus, Haemophilus, Fusobacterium*, previously noted in IPMN graded with HGD or cancer. These are highlighted in **Supplementary Figure 6-7 cluster 2**. Subsequent correlation analysis revealed that the increased level of bile acid metabolites were significantly attributed by mainly *Enterobacteriacea, Streptococcus, Pseudomonas*, while the TMAO was attributed by *Klebsiella* (p<0,05), while kynurenine in turn appears to be attributed by the *Rhobdobacter* (p<0,05) (**Supplementary Table 2)**.

**Figure 6.**
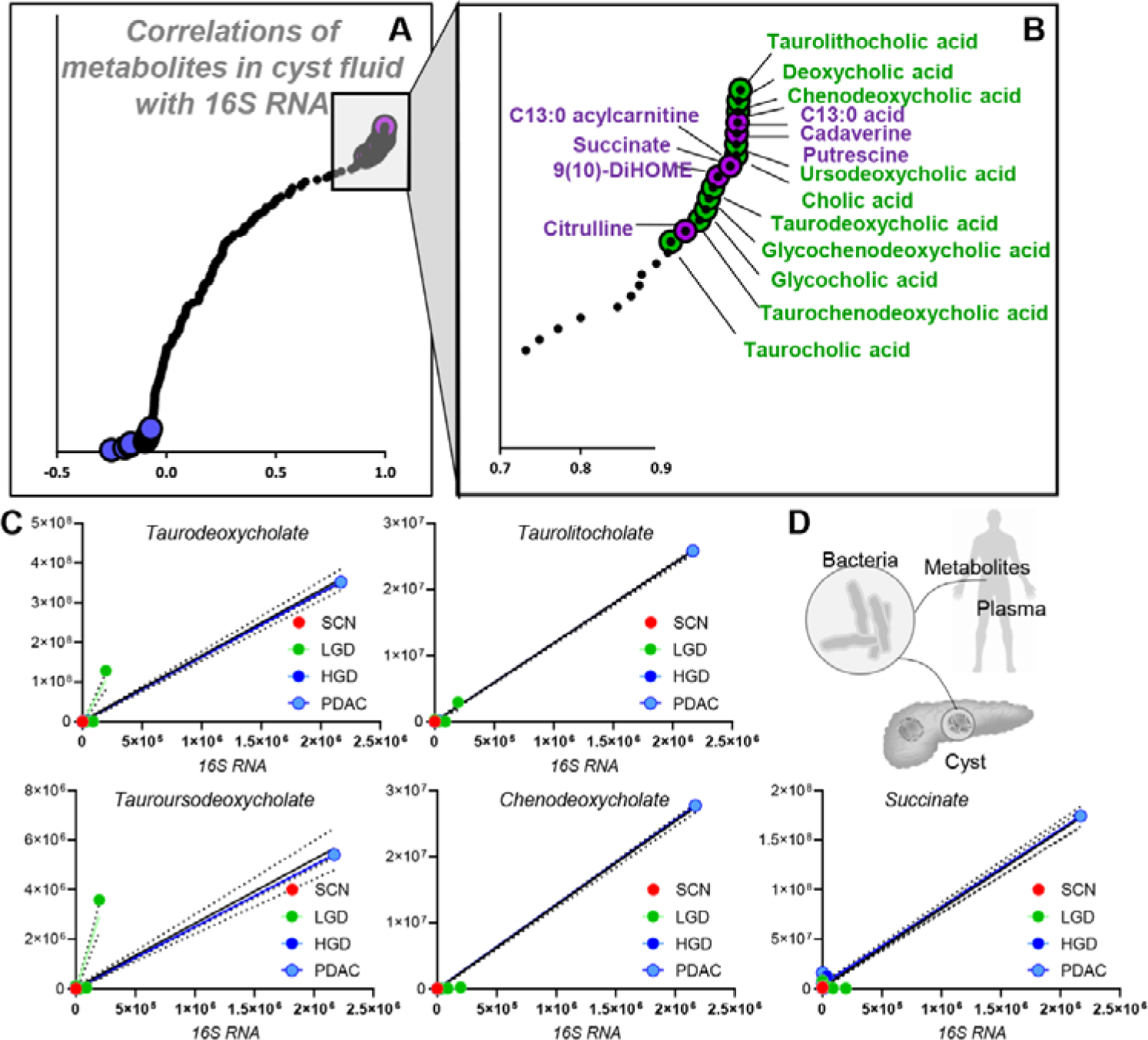
Correlation of 16S RNA data to cyst fluid and plasma metabolites in this study. **(A)**. Several carboxylic acids, polyamines and conjugated bile acids were identified among the top positive correlates to the total 16S RNA levels (**B-C**), suggestive of the model proposed in **D**.

**Figure 7.**
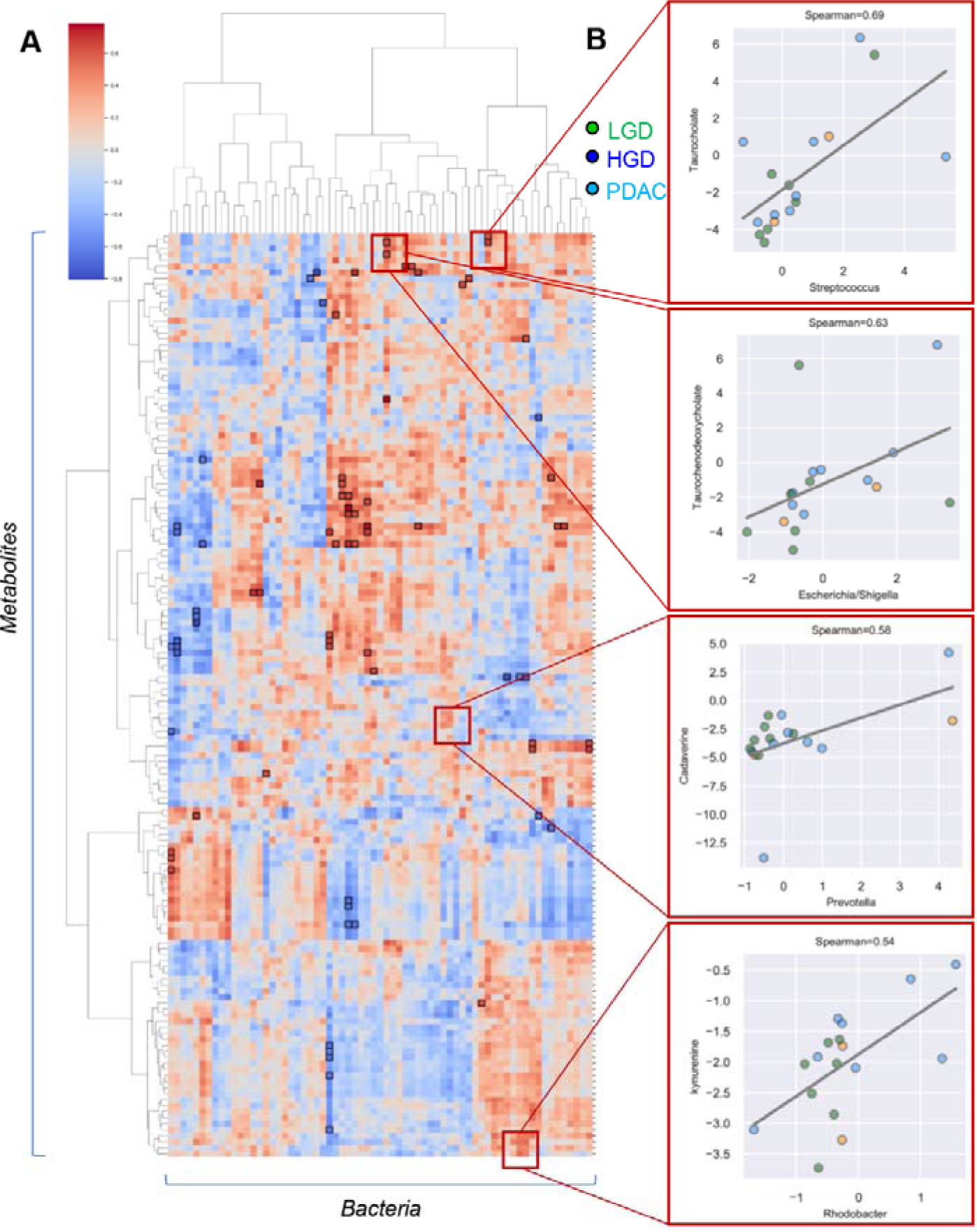
Hierarchical clustering of metabolic correlates to 16S RNA levels divided by bacterial strain. **(A)** identified clusters of metabolites and bacteria. A few selected line plots of significant correlates are highlighted in **B** as a function of IPMN status (grade of dysplasia, either low or high – LGD and HGD, respectively) or pancreatic ductal adenocarcinoma (PDAC).

## Discussion

While pancreatic IPMNs are common precancerous lesions – with incidence as high as ∼49% (9) – limited tools are currently available in the clinics to identify patients with elevated risk of cyst progression to cancer. Current approaches rely on radiological assessment of cyst diameter, pancreatic duct dilatation, rate of progression and clinical markers such as CA19-9, carcinoembryonic antigen and amylase (7). Unfortunately, these tools are either very specific, but not sensitive (e.g., CEA in malignant and invasive IPMN has >93% specificity but only 18% sensitivity (18)), or do not allow to determine IPMN-associated grade of dysplasia nor to provide a differential diagnosis for benign and low risk cysts, such as SCN. Recently, we have reported on the use of mass spectrometry-based metabolomics approaches as a tool to identify molecular markers of disease progression in patients presenting with IPMN cysts of different degree of dysplasia or pancreatic cancer prior to pancreas resection (11). As a result, we had identified both in pre-operative plasma and peri-operative cyst fluid a subset of candidate markers that could discriminate between IPMN and serous cystic neoplasm (SCN). In particular, amino acids were identified as the top discriminating molecules between the two groups in plasma and free fatty acids correlated significantly with the carbohydrate antigen CA19-9 (11). Of note, we (19) and others (20, 21) had also independently reported a role for the circulating levels of certain amino acids (e.g., lysine, histidine, proline) or lipids (sphingolipids and ceramides) to enable early detection of pancreatic adenocarcinoma. However, while our previous study managed to identify markers that separated SCN samples from the IPMN samples, unsupervised analyses showed a poor separation of the IPMN samples independently of the grade of dysplasia or pancreatic cancer stage (11). To overcome such limitations, in the present study we expanded on these preliminary observations by performing untargeted metabolomics and targeted quantitative measurements of selected metabolites to identify novel markers, while validating previously identified candidate markers in a newly enrolled independent cohort. As a result, we confirmed that several amino acids – including alanine, cystine, glutamine, histidine, lysine, ornithine, proline, serine, threonine - varied significantly across the various groups, but we also report significant alterations in the plasma and cyst fluid levels of several carboxylic acids (citrate, itaconate, 2-hydroxyglutarate, fumarate, malate), sugars and glycolytic metabolites (glucose, glyceraldehyde 3-phosphate, lactate), purine oxidation products (hypoxanthine, xanthine, urate and allantoate), heme metabolites (bilirubin and biliverdin) and free or carnitine-conjugated fatty acids. While in our previous study we had shown that free fatty acids correlated with cancerous state, as gleaned by the level of CA19-9, here we validated this observation and provide quantitative thresholds for several carnitine-conjugated fatty acids as discriminants between SCN and IPMN (all groups), as well as SCN or LGD cysts from more severe HGD and PDAC patients. Notably, while CA19-9 is a good marker of disease progression, studies have shown that this marker cannot discriminate between HGD and pancreatic cancer (18, 22). On the other hand, here we show that HGD and PDAC can be discriminated based on the plasma or cyst fluid levels of several amino acids (tryptophan) and related catabolites (monoamine oxidase products of tyrosine metabolism: adrenaline, dopamine) of potential bacterial origin (indole). Interestingly, short and odd chain fatty acids have long been associated with microbial metabolism (23). Similarly, another top marker, TMAO is a molecule generated from choline, betaine, and carnitine via gut microbial metabolism. Recently, we reported alterations of TMAO metabolism in response to impairments in iron metabolism (dietary or transfusion-related), as a result of gut dysbiosis adversely impacting proliferation of *Parabacteroides* and favouring infections by *Clostridia* (14). Here we also report that several conjugated bile acids ranked amongst the top 10 predictors of IPMN status and disease progression. Since catabolism of conjugated bile acids is performed by bacteria, accumulation of pro-inflammatory taurine and glycine-conjugated bile acids have been observed in the context of inflammation, obesity, trauma/haemorrhagic shock or iron-induced dysbiosis (14–17)). Also, bile acids have been recently reported to drive the maturation of newborn’s gut microbiota.(24) Altogether, these findings demonstrate a metabolic active proinflammatory pancreatic microbiota is supportive of malignant succession of IPMN, evidence corroborated by direct proofs of bacterial DNA in the peri-operative cyst fluids as determined by 16S gene sequencing, and by our unpublished finding that sequenced live bacteria are recovered from cyst fluid of HGD and PDAC cases (the latter in manuscript). Bacterial 16S rRNA gene sequence analysis here confirmed our previous reports about an enrichment in the cysts of certain strains (e.g., *Fusobacterium nucleatum* and *Granulicatella adiacens*) (12). In addition, here we report the frequent characterization of phyla such as *Proteobacteria, Actinobacteria*, followed by *Firmicutes* and *Alphaproteobacteria*. Total 16S bacterial 16S DNA levels in the cyst fluid correlated to disease progression (median levels increasing in IPMNs from LGD to HGD and PDAC). We thus identified metabolic correlates to 16S DNA levels, both as a total and as a function of enrichment of specific bacterial taxa in the cyst. As a result, we showed that the levels of several metabolites (conjugated bile acids, (odd chain) free and carnitine-conjugated fatty acids, carboxylic acids, glycolytic metabolites and purine oxidation products) can be broken down into two separate clusters of which one cluster were enriched by bacteria members previously noted to be enriched in IPMN graded with HGD or cancer (12) including *Enterobacteriacea, Granulicatella, Klebsiella. Stenotrophomonas, Streptococcus, Haemophilus, Fusobacterium*. Our notion that these bacteria members appear mostly positively associated with microbiota-related metabolites (bile acids, TMAO, cadaverine, putrescine, as well as choline and carnitine which are involved in conversion of TMAO, and tryptophan derived indole and kynurenine), indicative that these are metabolically active bacteria residing in the cystic precursors to invasive pancreatic cancer. This has important clinical implication as microbiota taxa residing within tumours have been found to confer tumour chemo-resistance due to microbial drug metabolism. Additionally, it also offers opportunities to model exposures to microbial products or microbial metabolites that influence cancer onset and progression.

In conclusion, in the present study we validated in a new independent cohort with orthogonal quantitative methods several markers of diseases progression in plasma and cyst fluid from SCN and IPMN (either LGD, HGD and PDAC). We thus quantified thresholds for these markers through the auxilium of stable isotope-labelled internal standards. Finally, we identified novel markers of IPMN status and disease progression including conjugated bile acids, free and carnitine-conjugated fatty acids and TMAO. We show that the levels of these metabolites of potential bacterial origin correlated with the degree of bacterial enrichment in the cyst, as determined by 16S gene analysis. Overall, our findings are interesting per se, owing to the validation of previous markers and identification of novel small molecule signatures of IPMN and disease progression. In addition, our findings further fuel the provoking debate as to whether bacterial infections may represent an etiological contributor to the development and severity of the disease in pancreatic cancer, in like fashion to other cancers (e.g., *Helicobacter pylori* and gastric cancer (25)). Future studies will determine whether live bacteria (including *Klebsiella, Granulicatella, Enterococcus*) can be recovered from surgically removed IPMN cysts, and whether these bacteria can be characterised *ex vivo* for genotoxin potential in pancreatic cell models.

## Methods

### Patient population, enrollment criteria and biofluid collection

Patients undergoing pancreatic surgery for suspected pancreatic cystic neoplasm (PCN) with post-surgically validated intraductal papillary mucinous neoplasm (IPMN) and serous cystic neoplasm (SCN) from 2016 to 2019 at Karolinska University Hospital, Sweden, were included. Excluded were cases without a cystic component, non-IPMNs/SCNs, or those without cyst fluid in the resected pancreas. This study follows the Helsinki convention and good clinical practice with permission of the Ethical Review Board Stockholm and the Karolinska Biobank Board (Dnr 2015/1580-31/1). Written informed consent was obtained from all patients.

### Pancreatic cyst fluid collection

Fresh resection specimens were received within 20□minutes of surgical removal and sampled by certified pathologists, in sterile conditions and on ice. Macroscopic assessment to identify the cystic lesion and main pancreatic duct was done by a specialist pancreatic pathologist. Fluid from the main pancreatic duct was collected using a syringe without needle. When the cystic lesion was readily identified in the intact specimen, the fluid was aspirated using a syringe with needle. For specimens in which the cystic lesion was not readily accessible from the surface the specimen was cut or when the cyst content was too viscous content was aspirated using a syringe without needle. Aspirated fluid was stored at −80□°C until further analysis.

### Peripheral blood collection and plasma isolation

Venous whole blood was collected in K2 EDTA Vacutainer® tubes (BD, Stockholm, Sweden) immediately prior to surgery. Within four hours of collection, blood was processed using Ficoll Paque Plus (GE Life Sciences, Uppsala, Sweden) gradient-density centrifugation following manufacturer’s instructions and the plasma fraction was stored at −80□°C until further analyses.

### Histopathological diagnosis and cyst fluid classification

Resection specimens were fixed in 4% formaldehyde and processed for routine histopathological diagnosis. The cystic lesions were classified by light microscopic examination of hematoxylin-eosin stained slides by a specialized pancreatic pathologist as IPMN or SCN. The grade of dysplasia in IPMN was assessed using a 2-grade (high/low) scale (henceforth HGD and LGD, respectively), according to current international standard (26). To make the cyst fluid classification more representative of the neoplastic epithelium that produces it, specimens showing <5% high-grade dysplasia (HGD) were classified as low-grade dysplasia (LGD). Specimens with concomitant invasive pancreatic ductal adenocarcinoma were classified as “PDAC” and considered as a separate class for further analyses. In total, 45 plasma samples (5 SCN, 20 LGD, 10 HGD, 10 PDAC) and 55 cyst fluid samples (5 SCN, 28 LGD, 8 HGD, 14 PDAC) were analyzed in this study.

### Sample processing and metabolite extraction

A volume of 10μl of plasma and 30 μl of cyst fluid was extracted either in methanol:acetonitrile:water (5:3:2, *v/v/v* – hydrophilic metabolites) or pure methanol (lipid extraction). After vortexing at 4°C for 30 min, extracts were separated from the protein pellet by centrifugation for 10 min at 10,000g at 4°C and stored at −80°C until analysis. For polar metabolites, half of the extracts were dried down via SpeedVac prior to resuspension in ddH2O + 0.1% formic acid, to facilitate ionization and chromatographic resolution of certain polar metabolites (e.g., betaine, kyurenine, etc.).(27, 28) For lipidomics analyses,(28) supernatants were diluted 1:1 (v/v) with 10 mM ammonium acetate for analysis by ultra-high pressure liquid chromatography coupled to mass spectrometry (UHPLC-MS). A total of 41 stable isotope-labeled internal standards for absolute quantitation were spiked in the lysis buffer, as extensively described in Supplementary Materials and Methods.

### Metabolomics

UHPLC-MS metabolomics analyses were performed as described,(27, 29) using a Vanquish UHPLC system coupled online to a high-resolution Q Exactive mass spectrometer (Thermo Fisher, Bremen, Germany). Samples were resolved over a Kinetex C18 column (2.1×150 mm, 1.7 µm; Phenomenex, Torrance, CA, USA) at 45°C. A volume of 10 μl of sample extracts was injected into the UHPLC-MS. Each sample was injected and run four times with two different chromatographic and MS conditions, as previously described (27) and detailed as follows: 1) using a 5 minute gradient at 450 µL/minute from 5-95% B (A: water/0.1% formic acid; B:acetonitrile/0.1% formic acid) and the MS was operated in positive mode and 2) using a 5 minute gradient at 450 µL/minute from 5-95% B (A: 5% acetonitrile, 95%water/1 mM ammonium acetate; B:95%acetonitrile/5% water, 1 mM ammonium acetate) and the MS was operated in negative ion mode. The UHPLC system was coupled online with a Q Exactive (Thermo, San Jose, CA, USA) scanning in Full MS mode at 70,000 resolution in the 60-900 m/z range, 4 kV spray voltage, 15 sheath gas and 5 auxiliary gas, operated in negative or positive ion mode (separate runs). These chromatographic and MS conditions were applied for both relative and targeted quantitative metabolomics measurements, with the differences that for the latter targeted quantitative post hoc analyses were performed on the basis of the stable isotope-labeled internal standards used as a reference quantitative measurement, as detailed below.

### Lipidomics

Samples were resolved as described (28) over an ACQUITY HSS T3 column (2.1 x 150 mm, 1.8 µm particle size (Waters, MA, USA) using an aqueous phase (A) of 25% acetonitrile and 5 mM ammonium acetate and a mobile phase (B) of 50% isopropanol, 45% acetonitrile and 5 mM ammonium acetate. Samples were eluted from the column using either the solvent gradient: 0-1 min 25% B and 0.3 mL/min; 1-2 min 25-50% B and 0.3 mL/min, 2-8 min 50-90% B and 0.3 mL/min, 8-10 min 90-99% B and 0.3 mL/min, 10-14 min hold at 99% B and 0.3 mL/min, 14-14.1 min 99-25% B 1and 0.3 mL/min, 14.1-16.9 min hold at 25% B and 0.4 mL/min, 16.9-17 min hold at 25% B and resume flow of 0.3 mL/min. isocratic elution of 5% B flowed at 250 µl/min and 25°C or a gradient from 0-5% B over 0.5 min; 5-95% B over 0.6 min, hold at 95% B for 1.65 min; 95-5% B over 0.25 min; hold at 5% B for 2 min, flowed at 450 µl/min and 35°C7. The Q Exactive mass spectrometer (Thermo Fisher Scientific, San Jose, CA, USA) was operated independently in positive or negative ion mode, scanning in Full MS mode (2 μscans) from 150 to 1500 m/z at 70,000 resolution, with 4 kV spray voltage, 45 sheath gas, 15 auxiliary gas.

### MS2 analyses for untargeted metabolomics

For discovery mode untargeted metabolomics, dd-MS2 was performed at 17,500 resolution, AGC target = 1e5, maximum IT = 50 ms, and stepped NCE of 25, 35 for positive mode, and 20, 24, and 28 for negative mode, as described in previous publications (30).

### Quality control and data processing

Calibration was performed prior to analysis using the Pierce™ Positive and Negative Ion Calibration Solutions (Thermo Fisher Scientific). Acquired data was then converted from .raw to .mzXML file format using Mass Matrix (Cleveland, OH, USA). Samples were analyzed in randomized order with a technical mixture (generated by mixing 5 μl of all samples tested in this study) injected every 10 runs to qualify instrument performance. This technical mixture was also injected three times per polarity mode and analyzed with the parameters above, except CID fragmentation was included for unknown compound identification (10 ppm error for both positive and negative ion mode searches for intact mass, 50 ppm error tolerance for fragments in MS2 analyses – further details about the database searched below).

### Metabolite assignment and relative quantitation

Metabolite assignments, isotopologue distributions, and correction for expected natural abundances of deuterium, 13C, and 15N isotopes were performed using MAVEN (Princeton, NJ, USA), (31) against an in house library of deuterated lipid standards (SPLASH® LIPIDOMIX® Mass Spec Standard, Avanti Lipids) and in house libraries of 3,000 unlabeled (MSMLS, IROATech, Bolton, MA, USA; IroaTech ; product A2574 by ApexBio; standard compounds for central carbon and nitrogen pathways from SIGMA Aldrich, St Louis, MO, USA) and 41 stable-isotope labeled internal standards (detailed below). Discovery mode analysis was performed with standard workflows using Compound Discoverer 2.1 SP1 (Thermo Fisher Scientific, San Jose, CA). From these analyses, metabolite IDs or unique chemical formulae were determined from high-resolution accurate intact mass, isotopic patterns, identification of eventual adducts (e.g., Na+ or K+, etc.) and MS2 fragmentation spectra against the KEGG pathway, HMDB, ChEBI, and ChEMBL databases. Additional untargeted lipidomics analyses were performed with the software LipidSearch (Thermo Fisher, Bremen, Germany).

### 16S RNA gene quantification and sequencing

Cyst fluid microbial copy number was measured by the 16S DNA qPCR assay, the microbiome composition and diversity by PacBio Single Molecule, full-length 16S rRNA gene sequencing (GATC Biotech, Konstanz, Germany), as previously described.(12)

### Statistical Analysis

Graphs and statistical analyses (either t-test or One-way ANOVA), as well as correlation analyses (Spearman) and calculation of Receiver Operating Characteristic (ROC) curves were prepared with GraphPad Prism 8.0 (GraphPad Software, Inc, La Jolla, CA), and MetaboAnalyst 4.0.(32) The same software was used to perform multivariate analysis, including principal component analysis, partial least square discriminant analysis and hierarchical clustering analyses. For the integrated microbiome and metabolome correlation analysis, the genus-level OTU table and the relative metabolite measurements of pancreatic cyst fluid samples were processed to include bacteria and metabolites that appear in a minimum of 3 or more patients. Zero counts were imputed with a pseudo-count of 0.001, normalized and clr transformed. The Spearman correlation was performed on the transformed data. For each pair, a p-value was calculated using a permutation test with 10,000 permutations. For data processing and statistical analysis, Python (Version 3.6.10) was used with the following packages: Pandas (Version 1.0.2) and NumPy (Version 1.18.1) for data processing, SciPy (Version 1.4.1) for the calculation of Spearman’s correlation and Matplotlib (Version 3.1.3) REF4 and Seaborn (Version 0.11.0) to visualize the data (33, 34).

### Targeted quantitative metabolomics

Targeted quantitative metabolomics analyses were performed on all plasma and cyst fluid samples against stable isotope-labeled internal standards (list, vendors and concentrations provided in **Supplementary Materials**). For this quantitative analysis, upon acquisition raw data were converted to .mzxml and peak areas for light and heavy (stable isotope-labeled) isotopologues were exported for each metabolite, as extensively described in prior methodological work (27, 28, 35). In details, absolute concentrations were determined using the following formula: [light] = (abundance light) / (abundance heavy) * [heavy] [dilution factor] where [light] = concentration of non-isotopically labeled (endogenous) metabolite, (abundance light) = total area abundance of endogenous metabolite, (abundance heavy) = total area abundance of isotopically-labeled standard, and [heavy] = known concentration isotopically-labeled internal standard.

## Data Availability

All the raw data and elaborations in this manuscript are provided in supplementary tables.

## Acknowledgments

The Mass Spectrometry Shared Resource of the University of Colorado School of Medicine is supported in part by funds from the Colorado Cancer Center Support Grant (P30CA046934) by the National Cancer Institute.

## Supplementary Figure legends

**Supplementary Figure 1 - Hierarchical clustering analysis of semi-targeted data highlighted changes in plasma levels of amino acids, carboxylic acids and purines**.

**Supplementary Figure 2 – ROC curves of top metabolic discriminant between cancerous and non-cancerous state from targeted absolute quant data**

**Supplementary Figure 3 - Hierarchical clustering analysis of semi-targeted data highlighted changes in cyst fluid metabolites between SCN and PDAC samples**. A few representative ROC curves for top discriminant metabolites are shown in the bottom half of the figure.

**Supplementary Figure 4 – ROC curves of top metabolic discriminants between high grade dysplasia (HGD) and PDAC samples**.

**Supplementary Figure 5 – Top 25 most common bacterial infiltrations in pancreatic cysts in the present study as determined by 16S RNA**.

**Supplementary Figure 6 – Metabolic correlates to bacterial strains as determined by 16S RNA levels (A) reveal two sub-clusters (B and C) of metabolites with strong correlation to infiltration of the cyst by specific bacterial strains**.

**Supplementary Figure 7 – Vectorial version of the map in Supplementary Figure 6**

**Supplementary Figure 8 – Detailed view of significant (bold font) metabolic correlates to specific bacterial strains (vectorial version of the map in figure 7.A)**.

**Supplementary Figure 9 - Highlighted line plots of top correlates metabolites vs bacteria**.

**Supplementary Table 1 – Raw metabolomics data from the analyses described in this study**

**Supplementary Table 2 – Metabolic correlates to bacterial strains as determined by 16S RNAs**.

